# COVID-19-related Fake News in Social Media

**DOI:** 10.1101/2020.07.06.20147066

**Authors:** Md. Sayeed Al-Zaman

**Affiliations:** Department of Journalism and Media Studies Jahangirnagar University, Savar, Dhaka, Bangladesh

**Keywords:** Fake news, social media: COVID-19, Fakebook, WhatsApp, public health, India

## Abstract

This study analyzes *N*=125 prominent fake news related to the COVID-19 pandemic spread in social media from 29 January to 11 April 2020. The five parameters of the analysis are themes, content types, sources, coverage, and intentions. First, the six major themes of fake news are health, religiopolitical, political, crime, entertainment, religious, and miscellaneous. Health-related fake news (67.2%) dominates the others. Second, the seven types of fake news contents have four main types: text, photo, audio and video, and three combined types: text & photo; text & video; and text & photo & video. More fake news takes the forms of text & video (47.2%), while the main types of content are less popular. Third, the two main sources of fake news are online media and mainstream media, where online-produced fake news (94.4%) prevails. Fourth, the main two types of coverages are international and national, and more fake news has an international connection (54.4%). Fifth, the intention of fake news has three types: positive, negative, and unknown. Most of the COVID-19-related fake news is negative (63.2%). Although fake news cases are unevenly distributed and repeatedly fluctuates during the period, a slow decrease of daily cases is noticed toward the end.

## Introduction

Fake news is a historical crisis of human communication that produces tension, disharmony, and misunderstanding in human society. In the age of digital communication and social networking, it gets a new momentum worldwide. We call it in many names: misinformation, disinformation, false news, inaccurate news, and rumor, though the problem remains almost the same. Amid the COVID-19 pandemic, the world is in grief with half a million dead. Health-related uncertainties give birth to a new phenomenon that is addressed as *infodemic* (a portmanteau of “information” and “epidemic”). In this situation, both true and false information galore. While true information help to mitigate the crisis, false information amplifies it. However, fake news propagation in social media recently catches the attention of researchers thanks to its prevalence irrespective of regions. While developed world with advanced technologies and tech-literates are getting control over fake news, India as a poor South Asian country and the largest democracy in the world has still been suffering from burgeoning online fake news. Diverse issues, such as healthcare, medication, disease, religion, and politics have also participated in this fake news crisis (Kadam & Atre, 2020). Amid this erratic situation, fake news in social media seems difficult to tackle. This research helps to understand the dynamic nature of fake news in social media related to the COVID-19 pandemic that might help to detect fake news in social media with less effort.

This study, having a focus on Indian social media fake news related to COVID-19, analyzes both digital and analog data to answer the research queries. The following discussion is divided into four main sections. The literature review discusses and summarizes the findings from previous relevant studies in this area. In the methodology section, the data collection and data analysis processes are presented in detail. The result section illustrates the major and minor findings of this study. Lastly, the discussion section presents the major findings based on relevant arguments and previous findings. Some limitations and strengths of this study are also discussed in this section.

## Literature Review

The outbreak of COVID-19 experiences another parallel epidemic led by fake news. It primarily disrupts public health communication and incites mass anxiety. Meanwhile, as a prompt response, a few studies have already appeared in the scholarly arena that deal with social media-produced fake news related to the COVID-19 pandemic. Some researches investigate the fake news propagation from a more behavioral, cultural, and sociolinguistic perspectives. For example, Rovetta & Bhagavathula (2020) analyze the attitudes of erroneous information and its consumption in Italy using Google Trends from January to March 2020. The result shows that the top scientific and COVID-19-related terms are “novel coronavirus,” “China coronavirus,” “COVID-19,” “2019-nCOV,” and “SARS-COV-2” along with top-five searches: “face masks,” “amuchina” (disinfectant), “symptoms of the novel coronavirus,” “health bulletin,” and “vaccines for coronavirus.” More interesting, fake news spread across the regions along with racism-related information. Laato et al. (2020) develop and test a research model to explore why social media users share fake news on COVID-19. Their results show that “a person’s trust in online information and perceived information overload are strong predictors of unverified information sharing.” (Laato et al., 2020, p. 1). Pennycook et al. (2020) in their two studies with over 1600 participants investigate why people believe and spread fake news about COVID-19. Their finding shows many people simply fail to determine the truth value of the news and share it while they fail to decide so. Erku et al. (2020) discuss three tendencies parallel to the COVID-19 pandemic: the growth of fake medicines, fake news, and medication false information. While fake news and fake medicines both have a detrimental impact on public health, they also emphasize on proper medications. These studies suggest that the COVID-19 pandemic deepens the fake news problem.

However, some studies state the opposite findings. For example, Casero-Ripolles (2020) in the secondary data analysis finds that people are actively consuming news that rises from 60% during pre-COVID-19-era to 92% during the COVID-19 pandemic. The study also shows that fake news detection capacity among people rises by 12% during the pandemic. Another interesting finding is that the more people consume news, the more they become capable to detect fake news. However, this study has two limitations: it only focuses on the United States, a country with advanced communication technology and media system, and its findings are contradictory (especially the second one) to other studies (Alvarez-Risco et al., 2020; Cinelli et al., 2020; Laato et al., 2020; Pennycook et al., 2020; Pulido Rodríguez et al., 2020; Rovetta & Bhagavathula, 2020) that find COVID-19 as an infodemic-maker. Also, both research trends do not give adequate attention and importance only to social media fake news.

A few researches are based on the comparative analysis of fake news across social media platforms. For example, Cinelli et al. (2020) analyze data from five social media platforms: Twitter, Instagram, YouTube, Reddit, and Gab, to explore the patterns of information diffusion about COVID-19. The result shows that all platforms have different volumes of false news and Gab is more susceptible to it. The spreading pattern of information related to COVID-19 is similar irrespective of platforms, according to the study. Interestingly, Gab is a popular social media platform for the far-right political activists and supporters. Therefore, it suggests fake news is more prominent among them than general users. In another study, Pulido Rodríguez et al. (2020) argues that social media users share fake news more than evidence-based news related to COVID-19, which is one of the primary threats to society. Well-distributed fake news in social media, according to them, leads to contradictory and poor decision making. Like Cinelli et al. (2020), this study also analyzes data from multiple social media platforms. Pulido Rodríguez et al. (2020) retrieved, classified, and compared 1923 fake and/or unverified posts related to COVID-19 from Twitter and Sina Weibo. The result shows that more fake news is published and shared on Twitter than on Sina Weibo. It is important to note that while others choose to use the term *fake news* to address the information-crisis amid the COVID-19 pandemic, Orso et al. (2020) propose to use “inaccurate news” instead of “fake news” to address most of the news with false information.

While the previous researches explore dynamic problems fake news pose in the COVID-10-era, some researches offer probable solutions to this crisis as well. Erku et al. (2020) think pharmacists are the key player in health communication. Therefore, to reduce medication false information, reliable information from the pharmacists should be distributed to the public and other health professionals. Naeem & Bhatti (2020) in their paper indicate how the COVID-19 pandemic causes an infodemic with a huge amount of false information. To battle this crisis, they suggest to prepare myth busters, fact-checkers, and credible sources relating to COVID-19. They further suggest to compile and circulate the necessary reliable information for the general public, students, and faculty to recognize fake news.

Political and legislative connections to fake news are also analyzed in several studies. Alvarez-Risco et al. (2020) address the COVID-19 as an occasion of infodemic that produces a high amount of social media-based fake news that hinders public health response in Latin America. In their study, they observe the fake-news-situation in Peru. Their observation states that the country’s strict law (i.e. imprisonment) against fake news production and propagation make it more successful in the battle. While Peru becomes more successful due to its governmental stringent step to tackle fake-news-problem, Dominican Republic struggles with dysfunctional governance (Tapia, 2020). The COVID-19 pandemic attacks the Caribbean country amid the political turmoil. Having a weak healthcare infrastructure along with public distrust, the government is losing its battle against the proliferating fake news epidemic. In another study, Gradon (2020) addresses the fake news problem from the perspective of Crime Science. It provides some preventive strategies to mitigate fake news propagation and proliferation: counter-narrative and use of AI as countermeasures are two of them. However, these studies do not consider fake news in social media, although their solutions might be applicable for controlling social media fake news as well.

Many studies offer possible solutions to social media-based fake news specifically. Although social media itself is a well-known producer of fake news, Orso et al. (2020) emphasize the positive use of social media to diffuse the necessary medical information amid this information crisis. They also suggest to filter social media communication to reduce the pervasiveness of *inaccurate* news, though it might sound undemocratic. Laato et al. (2020) also offer almost a similar solution to the problem. In another study, Pennycook et al. (2020) test an intervention intended to increase the truthfulness of the content people share on social media. They identify that sharing behavior improves after nudging the users to think about news accuracy. They further believe that *accuracy nudges* could be an immediate prevention against the tide of COVID-19-related fake news in social media. These studies mainly deal with social media-based fake news problem from a more behavioral, political, and preventive perspectives. However, no study addresses the necessity to identify the compact nature of COVID-19-related fake news that is produced, lived, and propagated in social media. To bridge this knowledge-gap this study tries to explore the necessary facets of social media-based fake news.

**RQ:** What are the prominent themes, content types, sources, coverage, and intentions of fake news in social media?

## Materials and Method

I trained and guided two graduate students to successfully perform their four research activities: (a) study the fake news in social media; (b) make a list of the fake news; (c) collect relevant data regarding the news, and (d) code the collected data. At first, they observed and searched social media for available fake news. In fake news searching and listing, they only focus on India, a fake-news-prone South Asian country, where social media penetration is unprecedented than the other neighbors (Kumar et al., 2013). After two weeks of continuous searching and studying fake news incidents, they listed 231 social media-based fake news from 29 January to 11 April 2020. (The first COVID-19 case was detected on 30 January 2020 and on 11 April 2020, the number of cases increased to 7529 with 242 dead, according to the Ministry of Health and Family Welfare of the Government of India.) The collected news was on various issues, such as religious, political, entertainment, and crime. After careful evaluation, they filtered out 106 news and finally, *N*=125 (54.11%) news remained. The primary filtering was based on two criteria set by the research question: (a) news that is unrelated to COVID-19; and (b) news that is absent in social media. The fake news meets the following five conditions as well: wide reach; wide acceptance; wide reactions; impact public and private life; proved as fake and debunked afterward; and present in social media platform(s). The students studied each of the news and tried to reveal their sources and contexts. However, many primary sources and contents of fake news were unavailable as they were removed after being debunked. In such cases, stored data (e.g., content and other relevant descriptions) were used. To seek stored data and other relevant information, we took help from an Indian fact-checking website, Alternative News (http://altnews.in). It works as a wing of the Pravda Media Foundation, a non-profit organization, and run by professional journalists and media specialists. The website is an archive of fake news across the country. Every article of this site debunks a particular fake news. One article includes background information of a case, link/mention of the fake news source, linked/uploaded contents of the fake news, details of the fake news: how it was produced and propagated, and on what grounds this story is fake. The website is highly credible for the data source and it is listed in the database of The Reporters’ Lab at Duke University managed by Bill Adair and Mark Stencel. Bill Adair is also the founder of PolitiFact (http://politifact.com), a Pulitzer prize-winning fact-checking non-profit website.

After the data collection, it was seen that fake news is more prevalent on four social media platforms: Facebook, WhatsApp, Twitter, and YouTube. The data analysis has two phases. One, the analysis of collected *contents* answered about the themes, content types, coverage, and intentions of fake news. Two, sources were detected either by *tracking*/*searching* the news or from the *descriptions* of the incidents in the articles. The collected fake news data must be two types: digital data from the internet, and analog data from the traditional media. Apart from analog media content (that often become popular in social media), user-generated content (UGC) from the internet are used widely in research purposes nowadays (Bordens & Abbott, 2017). Many of the previous literature used such contents as the primary data as well (Cinelli et al., 2020; Erku et al., 2020; Rovetta & Bhagavathula, 2020). In this quantitative content analysis, deductive coding of the collected data was performed. Two trained coders coded the collected data. They resolved the coding issues and complexities based on mutual consent and completed the codebook (Krippendorff, 2013). The coding was performed on the five parameters mentioned earlier. The whole process of collecting and processing data took almost a month (from mid-April to mid-May 2020) to complete.

## Results

The result shows that social media produces *N*=125 COVID-19-related fake news in a span of 84 days, from 29 January to 11 April 2020. The cases are unevenly distributed through the time. For example, the numbers of cases are likely to increase slightly after 27 February but drop after 10 March (see the Graph). The number rises again after 16 March and fluctuates constantly before reaching its peak on 27 March (*n*=9; 7.2%). The number drops starkly the next day, on 28 March (*n*=1; 0.8%) (Table 01). Before another heavy fluctuations, the number again reaches to the third-highest on 2 April (*n*=6; 4.8%). However, the curve shows a decline in daily cases from 9 April onward.

**Table 01:** Frequency distribution of fake news.

| Date | Frequency | Percent | Date | Frequency | Percent |
| --- | --- | --- | --- | --- | --- |
| 27-Mar-20 | 9 | 7.2 | 10-Apr-20 | 2 | 1.6 |
| 23-Mar-20 | 8 | 6.4 | 29-Jan-20 | 1 | 0.8 |
| 24-Mar-20 | 7 | 5.6 | 30-Jan-20 | 1 | 0.8 |
| 26-Mar-20 | 7 | 5.6 | 1-Feb-20 | 1 | 0.8 |
| 18-Mar-20 | 6 | 4.8 | 2-Feb-20 | 1 | 0.8 |
| 2-Apr-20 | 6 | 4.8 | 4-Feb-20 | 1 | 0.8 |
| 25-Mar-20 | 5 | 4.0 | 5-Feb-20 | 1 | 0.8 |
| 4-Apr-20 | 5 | 4.0 | 6-Feb-20 | 1 | 0.8 |
| 5-Apr-20 | 5 | 4.0 | 8-Feb-20 | 1 | 0.8 |
| 20-Mar-20 | 4 | 3.2 | 10-Feb-20 | 1 | 0.8 |
| 22-Mar-20 | 4 | 3.2 | 11-Feb-20 | 1 | 0.8 |
| 6-Apr-20 | 4 | 3.2 | 15-Feb-20 | 1 | 0.8 |
| 7-Feb-20 | 3 | 2.4 | 16-Feb-20 | 1 | 0.8 |
| 14-Mar-20 | 3 | 2.4 | 22-Feb-20 | 1 | 0.8 |
| 1-Apr-20 | 3 | 2.4 | 24-Feb-20 | 1 | 0.8 |
| 9-Apr-20 | 3 | 2.4 | 27-Feb-20 | 1 | 0.8 |
| 3-Feb-20 | 2 | 1.6 | 13-Mar-20 | 1 | 0.8 |
| 4-Mar-20 | 2 | 1.6 | 15-Mar-20 | 1 | 0.8 |
| 10-Mar-20 | 2 | 1.6 | 16-Mar-20 | 1 | 0.8 |
| 17-Mar-20 | 2 | 1.6 | 28-Mar-20 | 1 | 0.8 |
| 19-Mar-20 | 2 | 1.6 | 30-Mar-20 | 1 | 0.8 |
| 21-Mar-20 | 2 | 1.6 | 31-Mar-20 | 1 | 0.8 |
| 29-Mar-20 | 2 | 1.6 | 3-Apr-20 | 1 | 0.8 |
| 7-Apr-20 | 2 | 1.6 | 11-Apr-20 | 1 | 0.8 |
| 8-Apr-20 | 2 | 1.6 | Total | 125 | 100 |

Social media fake news related to COVID-19 has seven dominant themes: health, religiopolitical, political, crime, entertainment, religious, and miscellaneous. Health-related fake news has the highest prevalence (*n*=84; 67.2%) followed by religiopolitical fake news (*n*=21; 16.8%) with a huge gap in-between (Table 02). These two themes comprise 84% of the total fake news. Only *n*=2 (1.6%) miscellaneous fake news is found that is the lowest in the list. Health-related fake news has a few subtopics, such as medicine, medical and healthcare facilities, viral infection, doctor-patient issues, and quarantine. Religiopolitical and religious fake news has a subtle difference: religiopolitical fake news deals with the political aspects of religious fake news, while religious fake news is concerned about the spiritual and social aspects of religion.

**Table 02:** Themes of fake news.

|  | Frequency | Percent | Cumulative Percent |
| --- | --- | --- | --- |
| Health | 84 | 67.2 | 67.2 |
| Religiopolitical | 21 | 16.8 | 84.0 |
| Political | 9 | 7.2 | 91.2 |
| Crime | 3 | 2.4 | 93.6 |
| Entertainment | 3 | 2.4 | 96.0 |
| Religious | 3 | 2.4 | 98.4 |
| Miscellaneous | 2 | 1.6 | 100.0 |
| Total | 125 | 100.0 |  |

Fake news in social media has four primary categories: text; photo; audio; and video. However, fake news can take more than one form at a certain time. In that case, two or more primary contents constitute other combined categories. For example, political fake news can take the forms of both a text and a video simultaneously. Considering this, social media fake news is found in seven forms in total: text; photo; audio; video; text & photo; text & video; and text & photo & video. Text & video (*n*=59; 47.2%) is the most popular combination of social media fake news followed by another combination: text & photo (*n*=40; 32%) (Table 03). Both combinations constitute 79.2% of the total contents. Photo as a type has the lowest (*n*=1; 0.8%) instance in the list. Three other main content types are also less popular: text (*n*=14; 11.2%); audio (*n*= 6; 4.8%); and video (*n*=3; 2.4%).

**Table 03:**
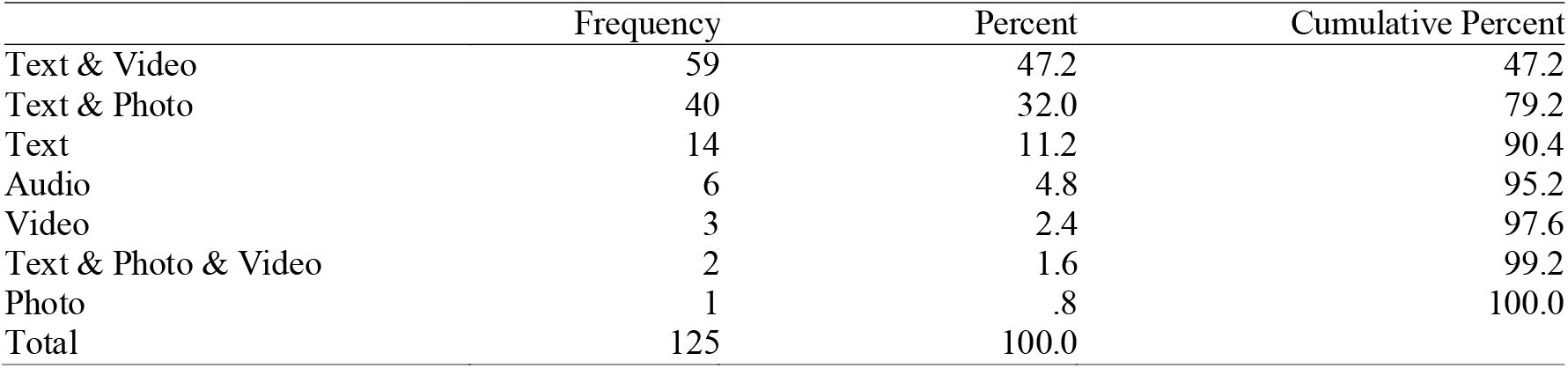
Types of Contents.

The two main sources of fake news in social media are online media and mainstream media. The mainstream media mainly includes television channels, newspapers, and radio channels. They are national media outlets. In contrast, online media includes online versions of mainstream television channels and newspapers, online news portals, blogs and websites, and social media platforms. Of the two types, online media (*n*=118) alone is responsible for 94.4% of the social media fake news, whereas mainstream media (*n*=7) is responsible for only 5.6% fake news. This uneven distribution suggests that online media is a far more effective fake news source amid COVID-19 than mainstream media. Of many sources of online media, four social media platforms are found producing the largest share of fake news: Facebook, WhatsApp, Twitter, and WhatsApp.

Relatively more social media fake news is international (*n*=68; 54.4%) than national (*n*=57; 45.6%) in nature and prevalence (Table 05). International fake news meets at least one of the three following criteria: globally-circulated fake news; has international connections/references; and talks about something international. In contrast, national fake news only deals with various domestic issues. Also, the intention of social media fake news related to COVID-19 is of three types: negative; positive; and unknown. Each fake news content is analyzed from one of the three aspects to determine its intention: the message the fake news conveys; the influences and impacts the fake news exerts; and the consequences the fake news produces. Thus, more fake news is found negative (*n*=79; 63.2%), whereas positive news is significantly lower (*n*=28; 22.4%) (Table 06). The intention of *n*=18 (14.4%) of fake news is undetected.

**Table 04:** Sources of fake news.

|  | Frequency | Percent | Cumulative Percent |
| --- | --- | --- | --- |
| Online media | 118 | 94.4 | 94.4 |
| Mainstream media | 7 | 5.6 | 100.0 |
| Total | 125 | 100.0 |  |

**Table 05:** Coverages of fake news.

|  | Frequency | Percent | Cumulative Percent |
| --- | --- | --- | --- |
| International | 68 | 54.4 | 54.4 |
| National | 57 | 45.6 | 100.0 |
| Total | 125 | 100.0 |  |

**Table 06:** Intentions of fake news.

|  | Frequency | Percent | Cumulative Percent |
| --- | --- | --- | --- |
| Negative | 79 | 63.2 | 63.2 |
| Positive | 28 | 22.4 | 85.6 |
| Unknown | 18 | 14.4 | 100.0 |
| Total | 125 | 100.0 |  |

## Discussion and Conclusion

The study seeks to explore the dynamic nature of fake news in social media that is related to COVID-19. A list of social media fake news from 29 January to 11 April 2020 was prepared. After filtering out, COVID-19-related *N*=125 fake news was compiled and relevant data and information related to each news were collected. I found that fake news is not consistent over time rather fluctuates, and a slow and subtle decrease in daily cases is noticed. It may support a result of Casero-Ripolles’s (2020) study that asserts news consumption during the pandemic increases remarkably; news consumption rate and fake news detection capacity have a positive correlation, which means the more the users consume news, the less the prevalence of fake news will be.

The prime aims of this study were to find out the themes, content types, sources, coverage, and intentions of the fake news cases. First, social media fake news related to COVID-19 has seven dominant themes: health, religiopolitical, political, crime, entertainment, religious, and miscellaneous. Of them, health-related fake news is on the top of the list that includes fake news mostly on medications and medical facilities. It supports a finding of Erku et al. (2020). However, the other crucial and sensitive fake news issues related to COVID-19 are ignored in the previous studies, such as religion, politics, and crime. Although Alvarez-Risco et al. (2020), Tapia (2020), and Gradon (2020) loosely discuss the connection between politics, crime, and fake news in different countries. Second, COVID-19-related fake news in social media has four primary content types along with three combinational types: text; photo; audio; video; text & photo; text & video; and text & photo & video. Interestingly, primary contents are less popular than their combinations. More fake news takes the forms of text & video followed by text & video: both are the combinations of three different primary contents and have a four-fifth share of the total percentage. No previous studies discuss the popular fake news content in social media related to COVID-19.

Third, the source identification of fake news in social media leads to an interesting finding. Almost all fake news is produced from online media except only a handful of cases that are from mainstream media. Also, four social media platforms: Facebook, WhatsApp, Twitter, and YouTube, contribute remarkably to the fake news production and dissemination. This result suggests how crucial online media can be in terms of fake news production and how important it is to guard online media to reduce the propagation and proliferation of COVID-19 fake news. Orso et al. (2020) and Laato et al. (2020) prescribe a similar solution for this problem, i.e. filter the social media news and communication. Fourth, people seem slightly more interested in international fake news than national fake news as more COVID-19-related fake news has an international connection, such as “Unrelated video viral as Italian police arresting man during COVID-19 lockdown” or “Dead bodies in Mecca shared as corona victims.” Such fake news has either international reference, or national issues having an international connection, or global reach. COVID-19 causes a global pandemic in this networked era; therefore, this result is expected. On the other hand, many COVID-19-related fake news in social media covers the national and local issues as well. Fifth, most COVID-19-related fake news is negative in nature, which can have detrimental impacts on the healthcare system and public health communication. Negative fake news conveys false information regarding medication, medical facilities, and death-related information that misguides both individual and community amid this uncertainty.

Apart from some exclusive findings, this study has a few limitations as well. The collected data are from Indian fake news cases and the country has a few distinctive natures from the aspects of political ambiance, cultural exceptionalities, technological penetration, communication, and news consumption patterns. These might elevate Indian fake news problems to another level that could be more relatable to Bangladesh and Pakistan, and more different from others, such as the USA and the UK (Al-Zaman, 2019, 2020). Also, the collected data are not cross-cultural and perhaps limited in number that might cause a generalization-problem. Moreover, a knowledge-gap still exists that has not been bridged yet: why COVID-19-related fake news in social media increases or decreases? After all its limitations, this study is a unique contribution to the COVID-19 fake news researches that identifies and bridges a few gaps in the existing literature. Besides, it explores further gaps that invite more researchers in this area.

## Data Availability

N/A

## Graph and Table

**Graph:**
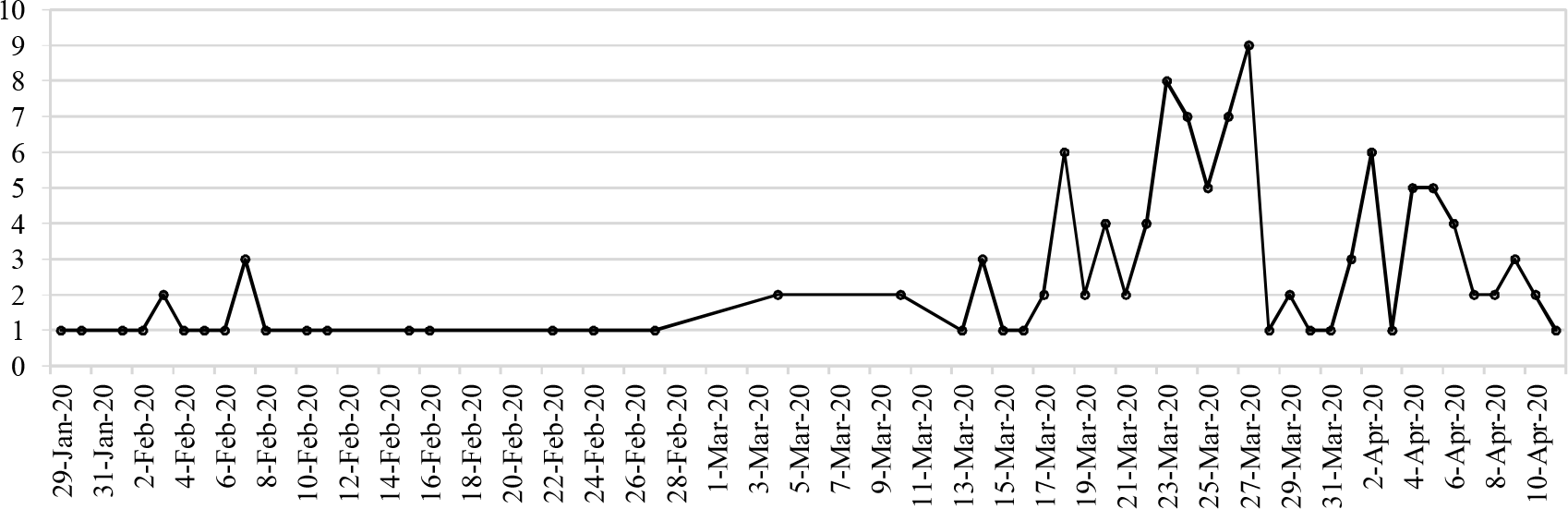
Timeline of fake news.

